# The COVID-19 Pandemic and Mental Health Concerns on Twitter in the United States

**DOI:** 10.1101/2021.08.23.21262489

**Authors:** Senqi Zhang, Li Sun, Daiwei Zhang, Pin Li, Yue Liu, Ajay Anand, Zidian Xie, Dongmei Li

**Affiliations:** Goergen Institute for Data Science, University of Rochester, Rochester, New York, USA; Department of Clinical & Translational Research, University of Rochester Medical Center, Rochester, NY, USA

**Author notes:** <u>Correspondence should be addressed to:</u> Dongmei Li, PhD, University of Rochester School of Medicine and Dentistry, Saunders Research Building 1.303J, 265 Crittenden Boulevard CU 420708, Rochester, NY 14642-0708, Zidian Xie, PhD, University of Rochester School of Medicine and Dentistry, Saunders Research Building 1.303D, 265 Crittenden Boulevard CU 420708, Rochester, NY 14642-0708. These two authors contributed equally to this study (Senqi Zhang, Li Sun).

**Keywords:** COVID-19, Mental health, Twitter

## Abstract

**Background:** Mental health illness is a growing problem in recent years. During the COVID-19 pandemic, mental health concerns (such as fear and loneliness) have been actively discussed on social media.

**Objective:** In this study, we aim to examine mental health discussions on Twitter during the COVID-19 pandemic in the United States and infer the demographic composition of Twitter users who had mental health concerns.

**Methods:** COVID-19 related tweets from March 5^th^, 2020 to January 31^st^, 2021 were collected through Twitter streaming API using COVID-19 related keywords (e.g., “corona”, “covid19”, “covid”). By further filtering using mental health keywords (e.g., “depress”, “failure”, “hopeless”), we extracted mental health-related tweets from the US. Topic modeling using the Latent Dirichlet Allocation model was conducted to monitor users’ discussions surrounding mental health concerns. Demographic inference using deep learning algorithms (including Face++ and Ethnicolr) was performed to infer the demographic composition of Twitter users who had mental health concerns during the COVID-19 pandemic.

**Results:** We observed a positive correlation between mental health concerns on Twitter and the COVID-19 pandemic in the US. Topic modeling showed that “stay-at-home”, “death poll” and “politics and policy” were the most popular topics in COVID-19 mental health tweets. Among Twitter users who had mental health concerns during the pandemic, Males, White, and 30-49 age group people were more likely to express mental health concerns. In addition, Twitter users from the east and west coast had more mental health concerns.

**Conclusions:** The COVID-19 pandemic has a significant impact on mental health concerns on Twitter in the US. Certain groups of people (such as Males, White) were more likely to have mental health concerns during the COVID-19 pandemic.

## Introduction

Coronavirus disease 2019, known as COVID-19, was first reported to be detected in China in December 2019. On March 13^th^, 2020, the declaration of a national emergency in the US marked the full outbreak of the pandemic. By July 5^th^, 2021, there were 30 million confirmed COVID-19 cases and 0.6 million related deaths in the US [1]. During this COVID-19 pandemic, the US people endured living in isolation and communicating in distance, and the country suffered from huge economic losses. The vaccine is a huge step towards the revitalization of lives. The US is striving to promote COVID-19 vaccines, and vaccinations are happening at an astounding rate. By June 30^th^, 2021, 3 billion vaccine doses have been administered worldwide [1]. Although a study pointed out that the pandemic will not end immediately even with the prevalence of vaccines [2], people’s physical health condition has greatly improved as vaccines offer protection to at least the degree of preventing severe diseases [3].

During the COVID-19 pandemic, there is another pressing issue - mental health conditions. In the US, 51.5 million adults have mental health issues according to the 2019 National Survey on Drug Use and Health data [4]. The cases of mental health illness are expected to drastically increase during the pandemic because of the restrictions, isolations, and sufferings. Many studies have discussed the impacts and consequences of the COVID-19 pandemic on mental health [5-7]. Studies found that COVID-19 sequelae include depression, anxiety, psychiatric disorder, and other mental health conditions [8, 9].

Previous studies have shown that social media is an ideal data source for studying mental health issues [10]. Other studies have taken mental health-related Twitter data into practical usage. For example, one study used Twitter data to build a model that can detect heightened interest in mental health topics [11]. Another study applied geographic information system (GIS) analysis on Twitter users who expressed depression [12]. During the COVID-19 pandemic, many studies emerged focusing on mental health topics using social media data. One study focused on tracking “loneliness” on one-month Twitter data [13]. Another study monitored the shift of mental-health-related topics and revealed the responsiveness of Twitter [14]. All those studies contributed greatly to raising awareness against mental health issues.

In this study, we tried to understand the mental health concerns during the COVID-19 pandemic in the US using Twitter data. Furthermore, we aimed to examine which demographic groups were most likely to have mental health concerns during the pandemic.

## Methods

### Data Collection and preprocessing

We used the Twitter streaming API to collect COVID-19 related Twitter posts (tweets) between March 5^th^, 2020 and January 31^st^, 2021 using COVID-19-related keywords, except from May 18^th^, 2020 to May 19^th^, 2020, and from August 24^th^, 2020 to September 14^th^, 2020 due to technical issues. The COVID-19 related keywords include abbreviations and aliases (“corona”, “covid19”, “covid”, “coronavirus”, “NCOV”) [15]. The dataset was filtered with health-related keywords from seven health-related categories including mental health, cardiovascular, respiratory, neurological, psychological, digestive, and other [16, 17]. There were 8,108,004 health-related tweets in the dataset. After removing duplicates, 8,044,576 tweets remained. To avoid the potential impact of promotion tweets, we filtered out the tweets that contained promotion-related keywords (“promo code”, “free shipping”, “percent off”, “% off”, “use the code”, “check us out”, “check it out”, “% discount”, “percent discount”). In this process, 4195 promotion-related tweets were removed. In our study, we focused solely on the mental health category.

Therefore, mental health-related keywords were used to derive a mental-health subset (“depression”, “depressed”, “depress”, “failure”, “hopeless”, “nervous”, “restless”, “tired”, “worthless”, “unrested”, “fatigue”, “irritable”, “stress”, “dysthymia”, “anxiety”, “adhd”, “loneliness”, “lonely”, “alone”, “boredom”, “boring”, “fear”, “worry”, “anger”, “confusion”, “insomnia”, “distress”) [17]. In this subset, there were 5,088,049 mental health-related tweets (Supplemental Figure 1).

To identify tweets from the United States, we further applied a geological filtering process to derive a US subset based on a US keywords list [18]. The US keywords list contained full names and the abbreviation of the country, states, and some major cities in each state. The filtering process was applied to the place of the tweets. Since most of the Twitter users preferred not to share the location for a single tweet [19], we continued the filtering on the “location” feature of users if “place” is empty. After this process, we derived our US mental health dataset, which contains a total of 1,270,218 tweets.

We used the “user_name” feature to identify the distinct users in the US mental health dataset. We examined the number of Twitter users posted their first mental health-related tweet and the number of mental health-related tweets they have posted during the study period. There were 591,022 distinct users in the dataset.

To study the relationship between the number of mental health-related tweets and daily COVID-19 cases in the US, we downloaded US daily case data from COVID Tracking Project (https://covidtracking.com/data/download) on March 19, 2021.

### Topic Modeling

The Latent Dirichlet Allocation (LDA) model was applied to extract the most frequent topics that people discussed relating to mental health during the pandemic. LDA is an unsupervised generative probabilistic model which, typically given the number of topics, allocates each word in a document to a specific topic, and calculates a weight for each word representing the probability of appearance in each topic [13]. First, we removed all punctuation, converted all texts to lowercase, and tokenized every sentence. Then, the Natural Language ToolKit package was applied to remove stop-words (e.g. the, is, a) [20]. Next, the Gensim package was applied to convert frequent bigrams and trigrams into a single term [21]. In this way, those phrases would be considered as one element in the modeling process. Lastly, we lemmatized all texts by converting all tenses to present tense and keeping only nouns, adjectives, verbs, and adverbs using spaCy [22]. We determined the optimal number of topics based on the coherence score, which measured the relative distances between keywords in each topic and the inter-topic distance map generated by LDAvis that visualized the overlap between topics [23]. We selected the topic number that had a relatively high coherence score.

### Demographic Inference of Twitter Users

We utilized a facial detection API provided by Face++ and a race/ethnicity prediction package called Ethnicolr to extract the demographic information of the users. Face++ is an AI open platform that applies deep learning to predict gender and age from an image [24]. Ethnicolr is a collection of several machine learning-based race and ethnicity classifiers trained on different data sets [25, 26].

First, we sorted the distinct user dataset based on the number of tweets per user posted in descending order. Since the average number of tweets per Twitter user posted is 2.15, we focused on 101,492 users who posted at least three mental health-related tweets. After downloading the profile image using the “profile_image_url” feature, we utilized the API to identify the number of faces, age, and gender in the image. Age and gender would be collected if the image only contains one face. As reported in a previous study [15], face++ has an accuracy of 93% in predicting gender. Age is much harder to be accurately determined, and the accuracy is 41%. To accommodate the situation, we choose to classify users into age groups to achieve better accuracy. We grouped the users into five age groups (<18, 18-29, 30-49, 50-64, ≥ 65) based on the criteria of the Pew Research Center [27]. We eliminated the 18-age group due to the small sample size (only 49 Twitter users).

We utilized the “census_ln” function in Ethnicolr, which trained on US census data in 2010 to predict race and ethnicity. It has an average accuracy of 79% on four races [16]. After removing all emoji and special characters, the algorithm takes a list of clean usernames with valid age and gender information as input to predict the probabilities of Non-Hispanic Whites, Non-Hispanic Blacks, Asians, and Hispanics for each name. We accepted the category with the highest probability as the prediction result. We obtained 11,330 Twitter users with valid age, gender, and race information.

## Results

### Longitudinal Trend of Tweets Related to Mental Health in the US

To understand how the COVID-19 pandemic might affect mental health in the United States over time, we performed a temporal analysis on the number of tweets mentioning mental health in the US. As shown in Figure 1, the number of mental health-related tweets fluctuated over time with three major peaks, including from late April to early May in 2020, middle June to late July in 2020, and late October to early November in 2020.

**Figure 1.**
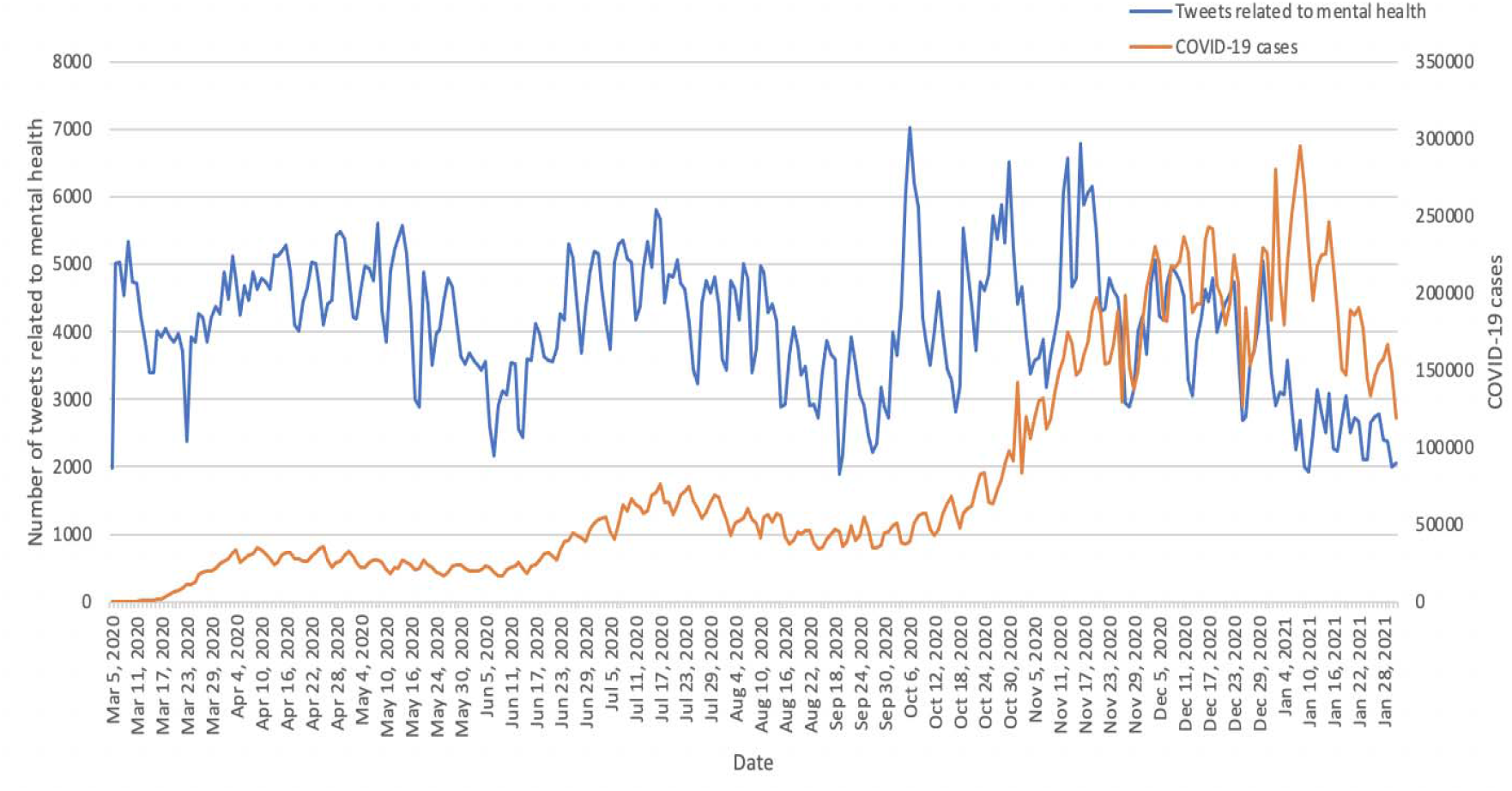
Number of COVID-19 tweets mentioning mental health and daily COVID-19 cases in the US.

On October 6^th^, 2020, there were 7,033 mental health-related tweets which was the highest number. To better understand the correlation between the number of mental health-related tweets and the severity of the COVID-19 pandemic, the number of daily COVID-19 cases in the US was shown in Figure 1. The Spearman’s correlation between the number of mental health tweets and the number of COVID-19 cases is 0.245 with p-value <.0001, which indicates that there is a mild positive correlation between the number of mental health-related tweets and the number of COVID-19 cases.

To examine which mental health keywords were the most mentioned on Twitter during the pandemic, we counted the appearances of each mental health keyword. After combining words that have the same meaning (For example, “depress” includes “depress”, “depression”, “depressed”), we divided the number of tweets for each keyword by the total number of tweets to calculate the proportion of each symptom. “Fear” was the most frequently mentioned symptom along with COVID-19, followed by “alone”, “failure”, and “depress” (Supplemental Figure 2).

### Major Topics Discussed in COVID-19 Tweets Mentioning Mental Health

To understand what might contribute to these mental health concerns mentioned in COVID-19 tweets, we performed topic modeling. As shown in Table 1, the first common topic is “stay-home and loneliness”, which has the highest percentage (24.70%) in all mental health-related tweets. The second topic is “death toll”, which is 16.70% of all tweets. The remaining topics have similar percentages, including “Politics and policy” (13.20%), “Personal symptom” (12.30%), “Covid and vaccine” (11.50%), “Help and relief” (10.90%), and “Government responses” (10.70%).

**Table 1.** Topics discussed in Tweets mentioning COVID-19 and mental health.

| Topic | % Token | Keywords | Examples |
| --- | --- | --- | --- |
| Stay-home and loneliness | 24.70% | covid, get, alone, tired, go, people, worry, mask, know, home | Joe, I am old... been **home alone** since mid-Feb. I have auto-immune issues. I have been sick 3 times this year after going to the market. I was very careful. I fear **Covid** and **worry** how long it will be for someone like me to get a vaccine. There are many like me. |
| Death toll | 16.70% | fear, covid, people, death, die, alone, virus, many, number, live | Yea but fearing it causes more damage than actual virus. Virus **deaths** is lie created to cause that **fear**. We can't **live** in **fear** always. I have been around a lot with covid and not once have caught it. **Covid** is not the big bad killer that dems and media want u to believe. |
| Politics and policy | 13.20% | covid, fear, biden, trump, country, vaccine, great, american, bill | I don't **fear Covid** specifically at all, though I should since I 'm high-risk. But yes, I do have that generalized anxiety. I think it's a combo of **Covid**, murder hornets, **Trump, Biden**, and riots in the streets. |
| Personal symptom | 12.30% | covid, fatigue, test, vaccine, get, depression, patient, hospital, symptom | Lost a job, couldn't attend my grandfather's funeral, may never get to see my other grandfather again as he has been given little time to live, have had to cope with severe **depression** in total isolation, actually had vivid, developed chronic **fatigue** and urticaria as a result... |
| Covid and vaccine | 11.50% | vaccine, covid, new, worry, alone, case, coronavirus, pandemic, day, worker | **Vaccines** offer protection against **COVID-19**, but experts **worry** side effects may scare people off |
| Help and relief | 10.90% | covid, stress, anxiety, pandemic, help, health, relief, mental, time | THIS VIDEO FROM @PrinceEa is going to help give **relief** from covid **anxiety** because you're being proactive and keeping in the best possible immune **health!** |
| Government responses | 10.70% | failure, covid, trump, american, response, coronavirus, dead, pandemic, lie, death | Literally what rules? This is **Failure** of Leadership. Slow to close and quick to reopen, like **trump wanted**. Require Masks. Get Money to Citizens for Rent, Food, Utilities. Close Bars + Restaurants. Ban Large Gatherings. This doesn't take a rocket scientist. just a leader |

### Twitter Users Who had Mental Health Concerns Related to COVID-19

To analyze how the COVID-19 pandemic affects the public on mental health over time, we examined the Twitter users who posted their first mental health-related tweet and calculated the number of unique Twitter users who posted their first tweet each day (Figure 2). Even though the number of Twitter users who posted their first mental health tweet varied over time, the overall number of new users who had mental health concerns was decreasing. On March 6^th^, 2020, there were 4,451 Twitter users who posted their first tweet related to COVID-19 and mental health, while there were no more than 1,000 new Twitter users in each day in January 2021. In addition, we examined the number of Twitter users who mentioned mental health each day during the pandemic (Supplemental Figure 3), which showed a similar trend as the number of mental health-related tweets (Figure 1).

**Figure 2.**
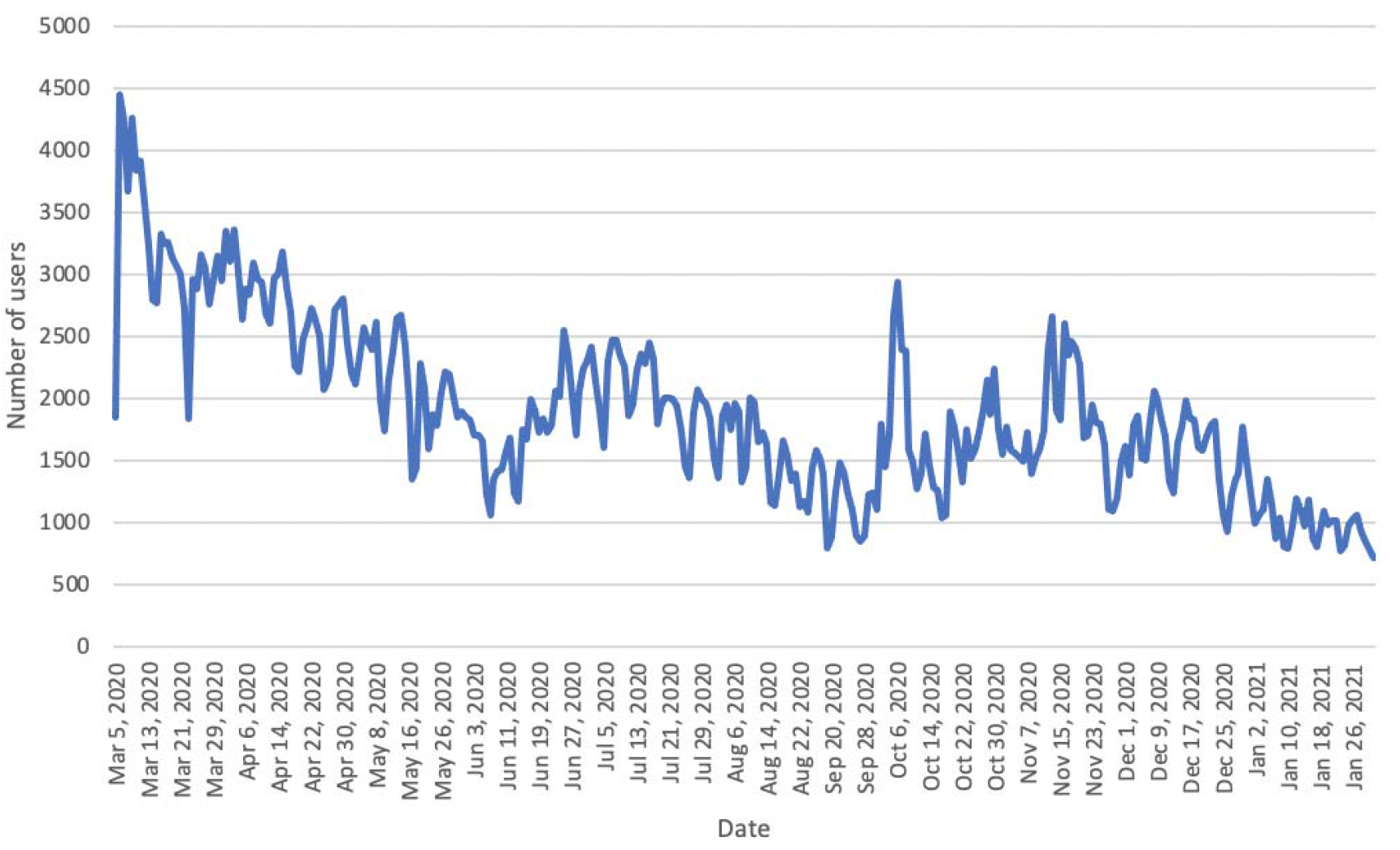
The number of US Twitter users having their first tweets about mental health over time.

### Geographic Distribution of Twitter Users Who had Mental Health Concerns in the US

While we have shown that the number of Twitter users who had mental health concerns in the US was large, it is important to examine whether there were some geographic differences in these Twitter users. To address this, we calculated the proportion of distinct Twitter users who mentioned mental health in each state, which was normalized by the state population. As shown in Figure 3, the states with a high proportion of Twitter users who mentioned mental health were centered to the east and west coast, such as Washington (359 Twitter users per 100,000 people), New York (244 Twitter users per 100,000 people), and Maine (471 Twitter users per 100,000 people). In addition, we examined the average number of mental health-related tweets per Twitter user in different US states, which showed that the states in the middle west (such as South Dakota) had a higher average number of mental health tweets per user (Supplemental Figure 4).

**Figure 3.**
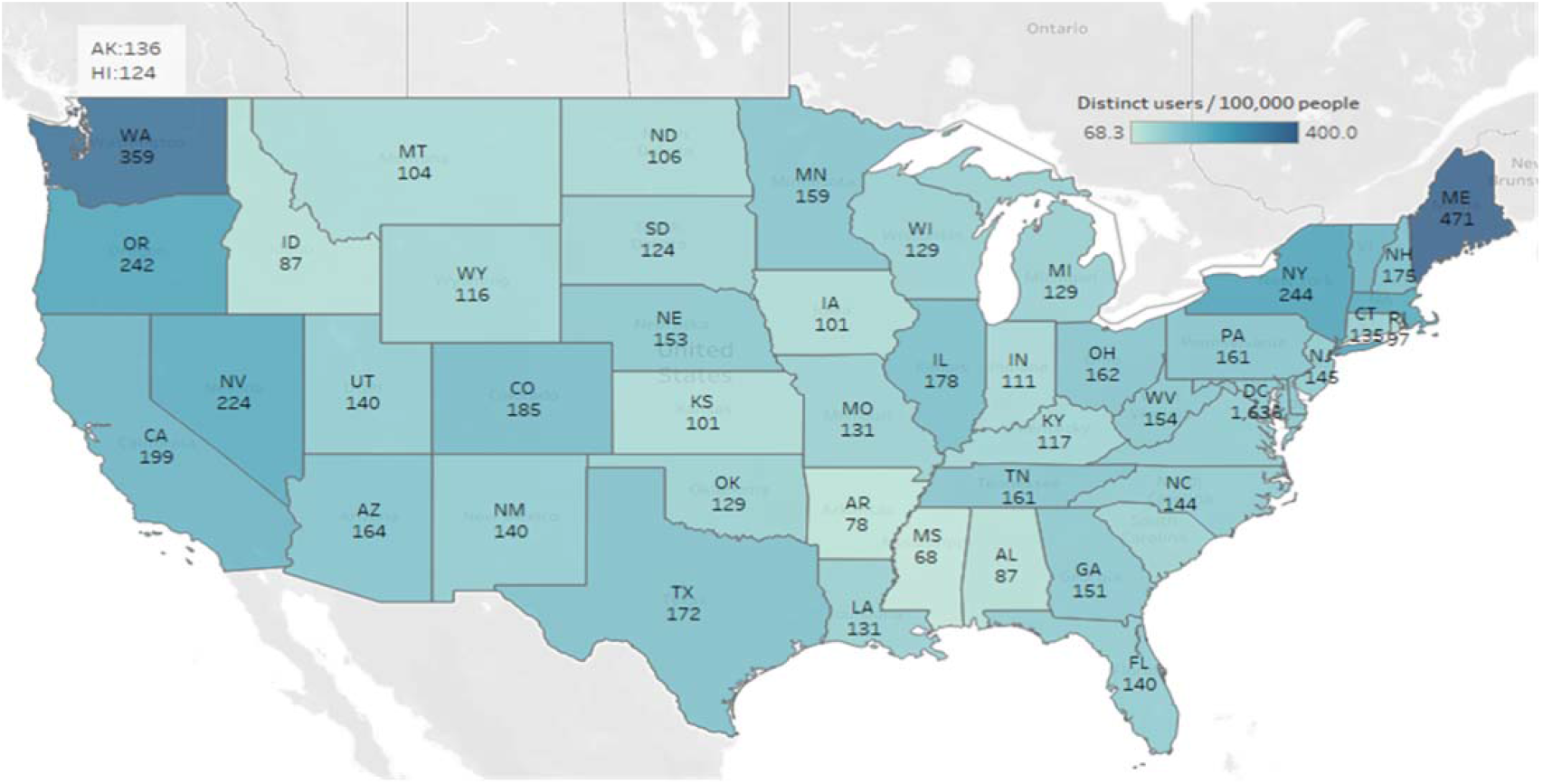
The proportion of Twitter users who had mental health concerns in different US states normalized by the population.

### Demographic Characteristics of Twitter Users Who had Mental Health Concerns in the US

To better understand the demographic composition of Twitter users, especially those who posted several mental health-related posts during the pandemic, we estimated thei demographic information using deep learning algorithms including Face++ API and Ethnicolr. As shown in Figure 4, 58.41% of Twitter users who were concerned about mental health were males while 41.59% were females. The age 30-49 group has the highest percentage (40.81%), followed by age 50-64 (30.10%), age 18-29 (15.38%), age 65 and above (13.28%). For race/ethnicity, White was the most (85.28%), followed by Asian (7.06%), Hispanic (5.25%), and Black (2.42%).

**Figure 4.**
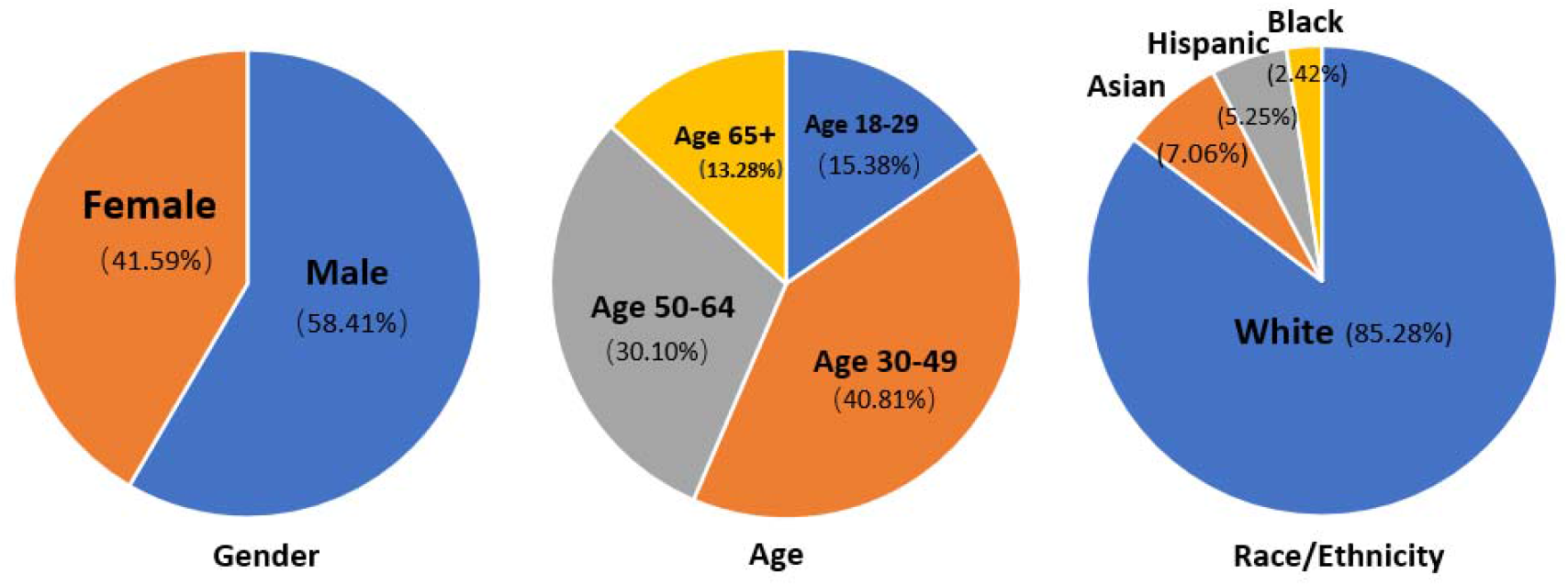
Demographic composition (including age, gender, and race/ethnicity) of Twitter users who had mental health concerns in the US.

By examining the gender distribution in each age group (Supplemental Figure 5), we showed that at the young age group (age 18-29), the proportion of females is significantly higher than the proportion of males (P<.0001). However, in the middle and old age groups (age 30-49, age 50-64, and age 65+), the proportion of males is significantly higher than that of females (P=.032, P=.0048, and P=.013 respectively).

Furthermore, we compared the proportion of each age group in each race/ethnicity (Supplemental Figure 6). By comparison, the proportion of Twitter users aged 30-49 with mental health concerns in the Asian group is significantly higher than that in the Black group (P=.0014) and White Twitter users (P<.0001). The proportion of Twitter users aged 18-29 with mental health concerns in the Asian group is significantly higher than that in the White group (P<.0001). White has a significantly higher proportion of Twitter users aged 50-64 with mental health concerns than Asian (P<.0001), Black (P = 0.026), and Hispanic (P=0.0058). The proportion of Twitter users aged 65+ with mental health concerns in the White group is significantly higher than that in the Asian group (P<.0001) and in the Hispanic group (P=.00046).

## Discussion

### Principal Findings

In this study, we observed a variation in mental health-related tweets over time and identified a moderate positive correlation between the number of mental health tweets and COVID-19 cases in the US, which suggests that the COVID-19 pandemic leads to more mental health concerns. We observed a downward trend of mental health-related tweets at the end of 2020 while the number of COVID-19 cases was still very high, which might be due to the success of COVID-19 vaccine development and the start of COVID-19 vaccination. Through keyword search and topic modeling, we identified the most pressing mental health concerns during the pandemic and related topics. The social distancing seemed to have severe effects on mental health concerns: people were forced to stay at home for a long time and traveling became risky and prohibited, which naturally led to feelings like stress and loneliness. For fear and anxiety, we infer that they came from the uncertainty about how long the pandemic will last and the high COVID infection and death rate. During the pandemic, it seemed that the public was not satisfied with some government responses and related health precaution policies, which might lead to the high frequency of “failure”.

In our demographic analysis, we estimated the demographic composition of Twitter users mentioning COVID-19 and mental health during the pandemic. Compared to the proportion of males in general US Twitter users (53.19%) [27], more male Twitter users (58.41%) had mental health concerns. Therefore, the males were more likely to express mental health concerns on Twitter. In the US, the proportion of people using Twitter decreases as the age increases (44.68% aged 18-29, 28.72% aged 30-49, 19.15% aged 50-64, 7.45% aged 65 or older) [27]. However, the majority of people posting mental health-related tweets during the pandemic were middle-aged and old-aged people (40.81% aged 30-49, 30.10% aged 50-64). The results indicate that middle-aged and old-aged people are more likely to express mental health concerns than young people on Twitter. Among all age groups, males were more likely to express mental health concerns except in the 18-29 age group. Compared with the distribution of the general Twitter users provided by a previous study [24], the proportion of White in Twitter users who mentioned mental health issues (85.28%) is significantly higher than the proportion of White in general US Twitter users (68%). However, the proportion of Asian (7.04%) and Black (2.14%) mentioning mental health issues is significantly lower than the proportion of general US Twitter users with Asian (18%) and Black (14%).

### Comparison with Prior Work

Mental health is one of the major health issues during the COVID-19 pandemic. One study conducted two rounds of surveys to investigate psychological impacts on people during the early phase of the COVID-19 pandemic in China [28]. Another study utilized an online survey and a gender-based approach to study the impact of the COVID-19 pandemic on mental health in Spain [29]. Both studies showed that the COVID-19 pandemic has a significant impact on mental health in public. In this study, we showed a positive correlation between the COVID-19 pandemic and mental health concerns on Twitter in the US.

Social media data had been used to study mental health issues during the COVID-19 pandemic. One previous study applied machine learning models to track the level of stress, anxiety, and loneliness during 2019 and 2020 using Twitter data [30]. The results showed that all of the three mental health problems (stress, anxiety, and loneliness) increased in 2020. Another study developed a transformer-based model to monitor the depression trend using Twitter [31]. The results showed that there was a significant increase in depression signals when the topic is related to COVID-19. These results are consistent with our results that stress, anxiety, loneliness, and depression are the top mentioned mental health emotions during the pandemic. Our topic modeling results further showed that loneliness is related to the quarantine at home.

While it is important to examine the impact of the COVID-19 pandemic on mental health, it might be more important to understand who might be affected the most by the pandemic in terms of mental health. One study applied the M3 (multimodal, multilingual, and multi-attribute) model to extract the age and gender information of Twitter users who posted COVID-19 related tweets from August 7 to 12, 2020, which showed that males and older people discussed more on COVID-19 and expressed more fear and depression emotion [32]. Another survey study showed that women and young people were more likely to have mental health issues and developed worse mental health outcomes during the pandemic [28]. In this study, we showed that there were more males, middle-aged and old-aged people discussing mental health-related topics on Twitter during the pandemic in the US. Besides gender and age, our study also estimated race/ethnicity information for Twitter users who tweeted about mental health during the pandemic, which provides a more comprehensive picture of the demographic portfolios of Twitter users having mental health concerns during the pandemic.

### Limitations

In this study, the mental health concerns on Twitter during the COVID-19 pandemic do not necessarily mean that these Twitter users had a mental illness. The keywords that we used for mental health concerns are relatively limited, which might introduce some biases. Another limitation lies in our demographic analysis. In our study, only a small proportion of users shared a valid human face as their profile pictures and a valid name. Of the 101,481 Twitter users we used for inference, only 11,330 (11%) users have valid names and profile pictures. Even if it’s a valid user, there is no guarantee that they are using photos and names of their own. In addition, due to the technical issue, we failed to collect the relevant Twitter data from May 18^th^, 2020 to May 19^th^, 2020, and from August 24^th^, 2020 to September 14^th^, 2020. Therefore, our data did not represent the whole population in the US.

## Conclusions and implications

During the COVID-19 pandemic, social media is one of the most popular platforms for the public to share their feelings. Our study successfully monitored the discussions surrounding mental health during the pandemic. As these topics revealed some causes of mental anxiety, they provided some directions for where efforts should be put to reassure confidence in the people. Our demographic analysis implicated that White and Males are more likely to have/express mental health concerns. Thus, more attention could be provided to them when mental health support becomes available. Furthermore, our study demonstrated the potential of social media data in studying mental health issues.

## Data Availability

The data are publically available from Twitter and can be accessed by anyone with internet connections.

## Acknowledgements

This study was supported by the University of Rochester CTSA award number UL1 TR002001 from the National Center for Advancing Translational Sciences of the National Institutes of Health. The content is solely the responsibility of the authors and does not necessarily represent the official views of the National Institutes of Health.

---

**Supplemental Figure 1.**
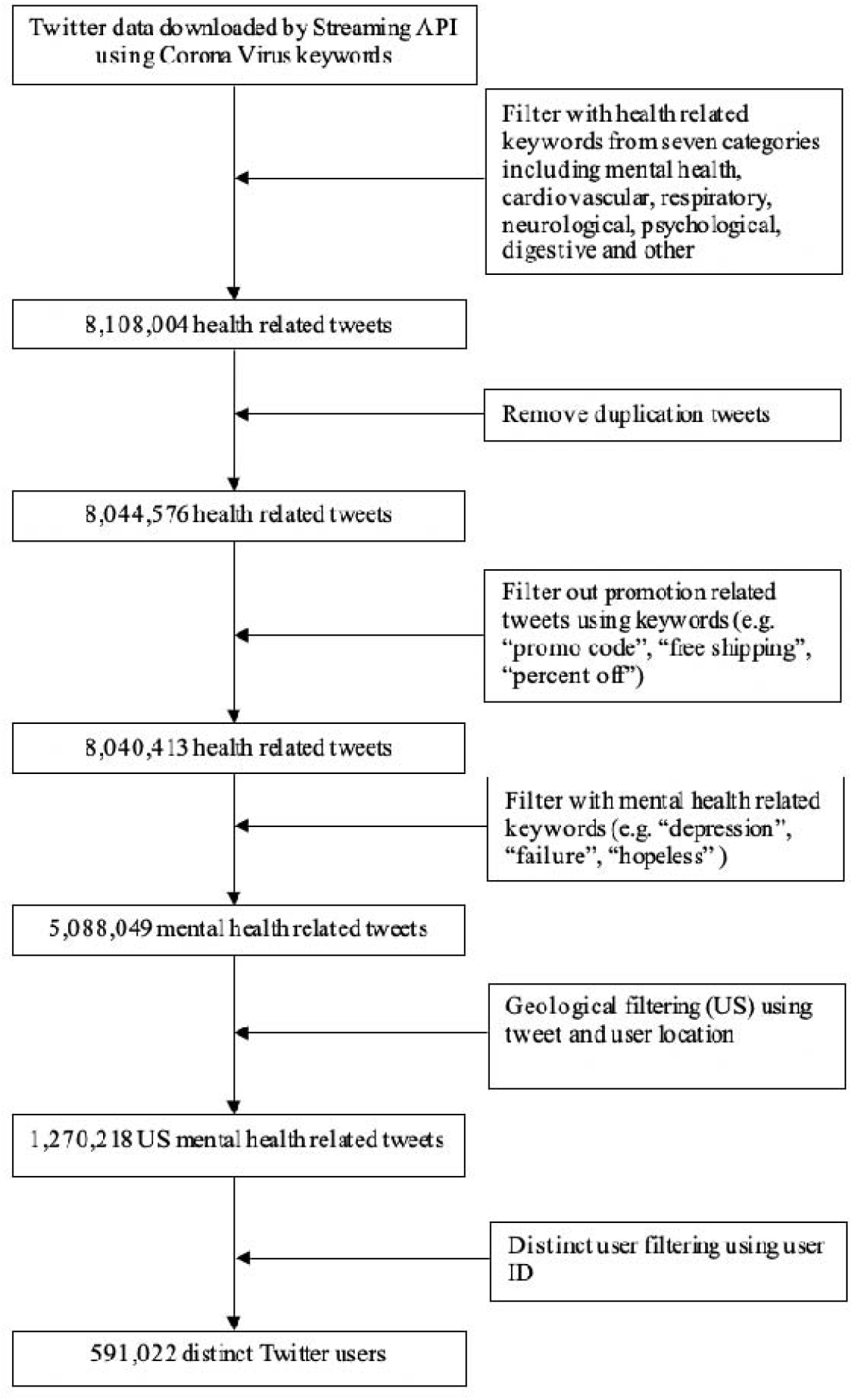
Flow chart of data pre-processing.

**Supplemental Figure 2.**
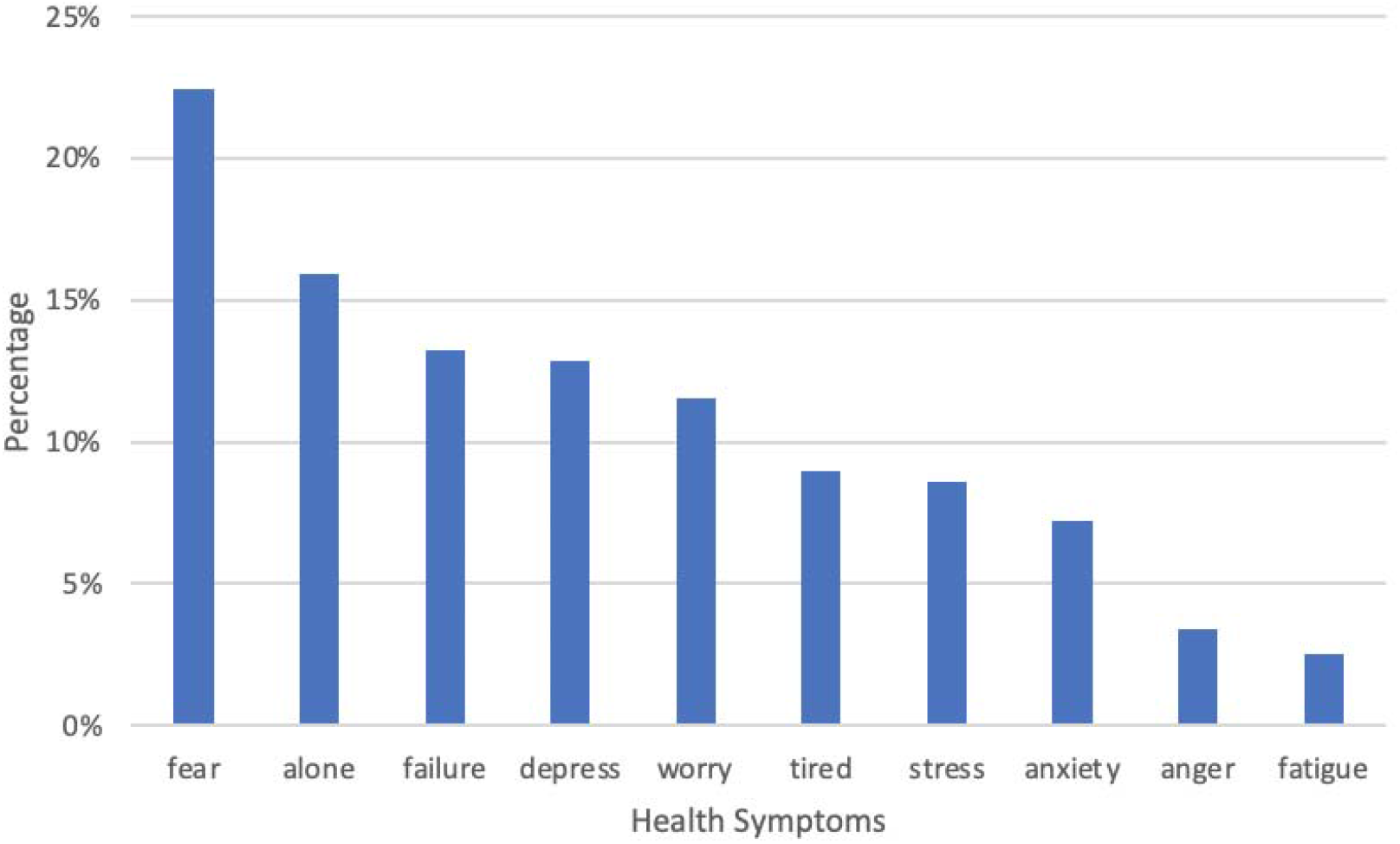
Mental health related keywords that were mentioned the most in COVID-19 related tweets in the US.

**Supplemental Figure 3.**
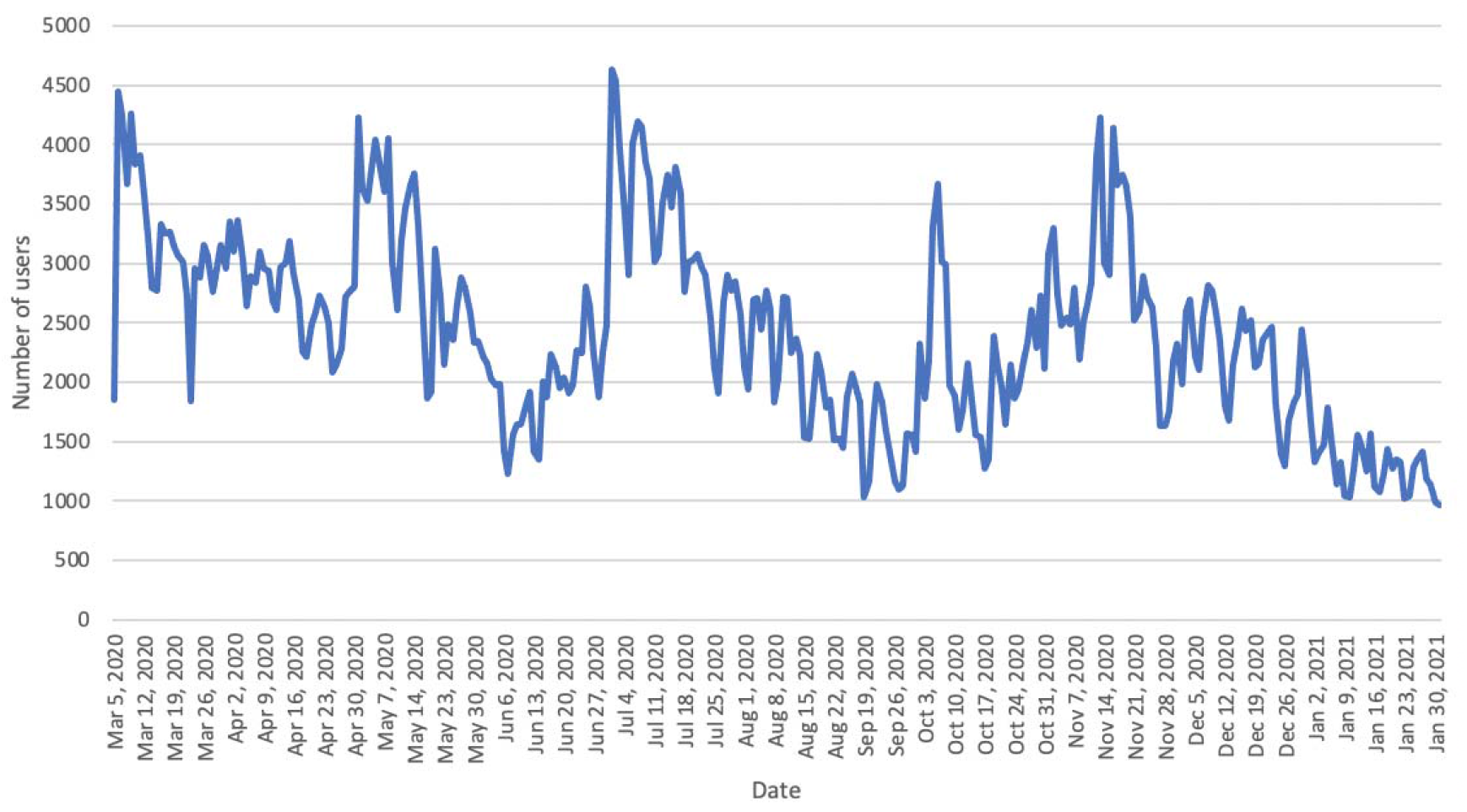
Number of Twitter users who had mental health concerns over time in the US.

**Supplemental Figure 4.**
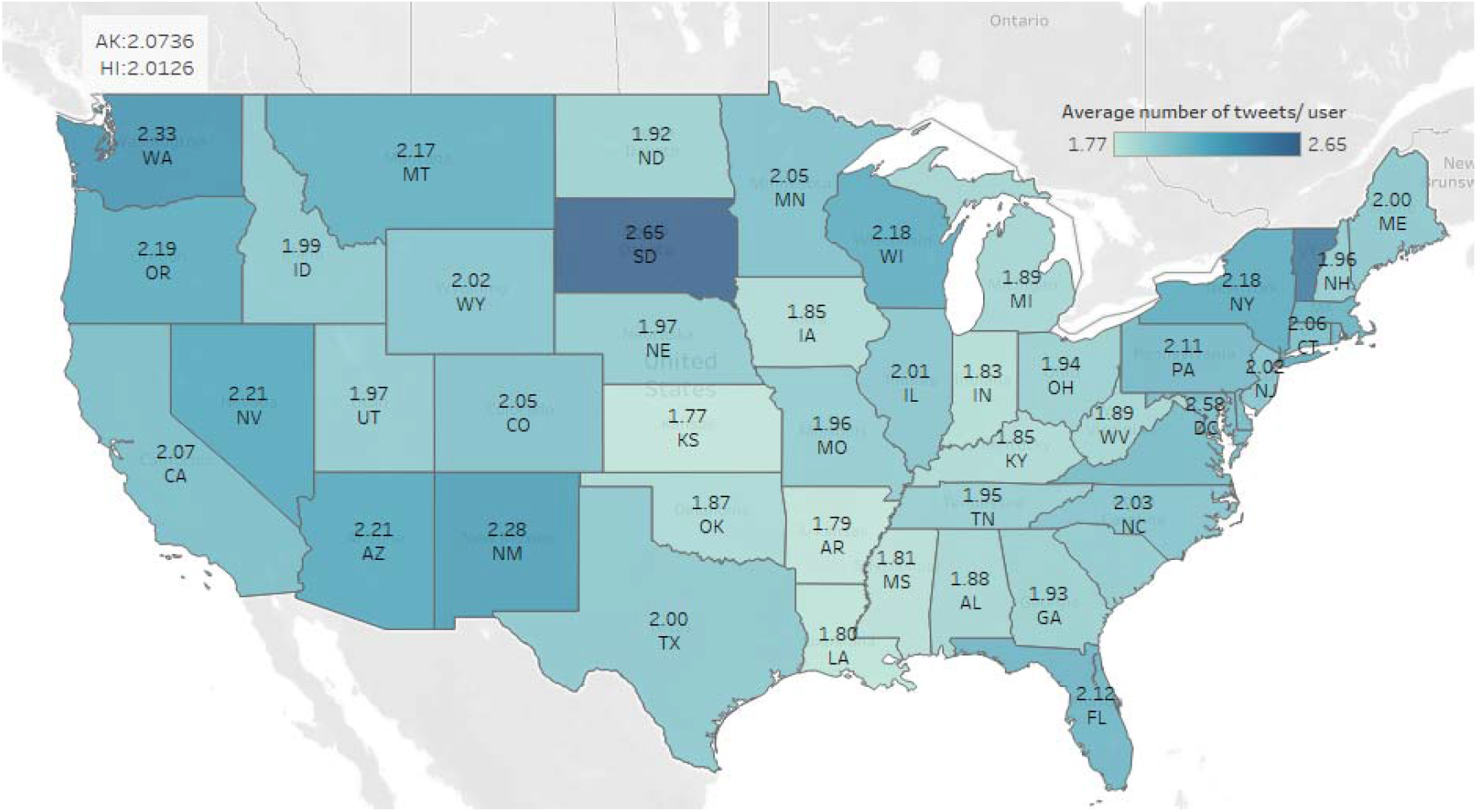
Average number of mental health related tweets per user in different US states.

**Supplemental Figure 5.**
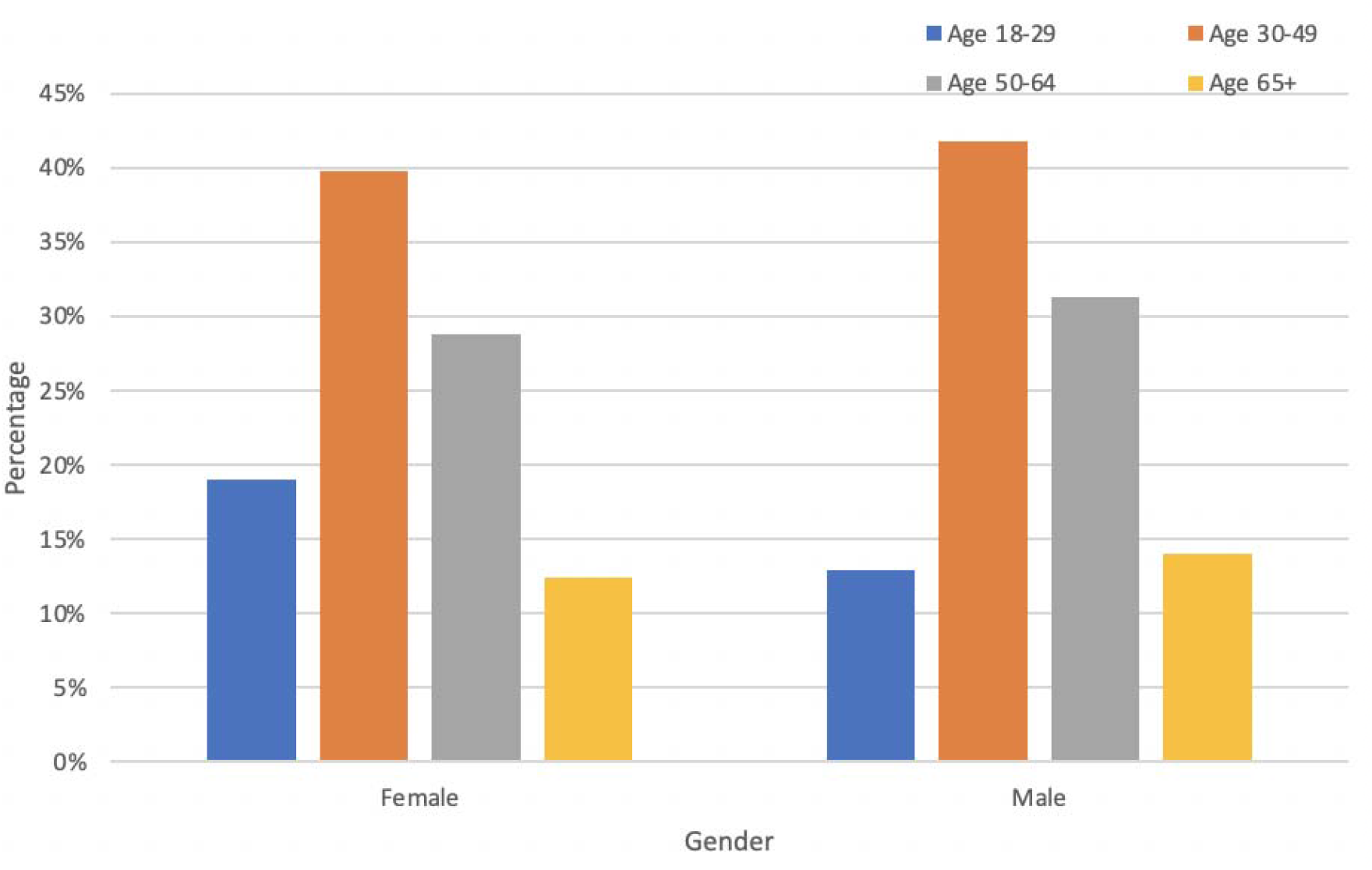
Age composition in different gender groups for Twitter users who had mental health concerns in the US.

**Supplemental Figure 6.**
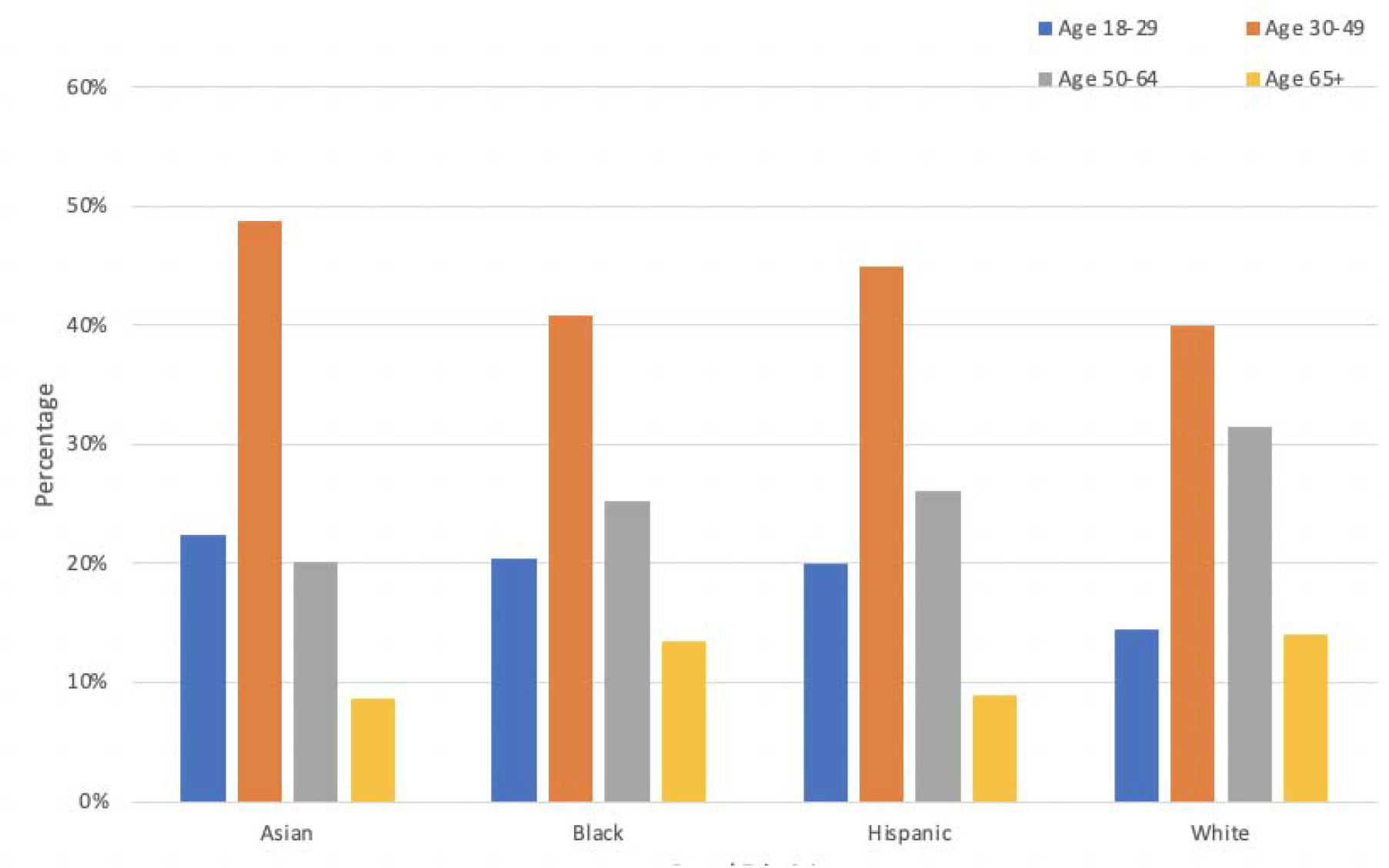
Age composition in different race/ethnicity groups for Twitter users who had mental health concerns on Twitter in the US.

## References

1. Covid-19 Data in Motion. 2021; Available from: https://coronavirus.jhu.edu/.

2. The Lancet, M., COVID-19 vaccines: the pandemic will not end overnight. The Lancet. Microbe, 2021. 2(1): p. e1–e1.

3. Katella, K. Comparing the COVID-19 Vaccines: How Are They Different? 2021; Available from: https://www.yalemedicine.org/news/covid-19-vaccine-comparison.

4. Mental Illness. 2021; Available from: https://www.nimh.nih.gov/health/statistics/mental-illness.

5. Pfefferbaum, B. and C.S. North, Mental Health and the Covid-19 Pandemic. New England Journal of Medicine, 2020. 383(6): p. 510–512.

6. Usher, K., J. Durkin, and N. Bhullar, The COVID-19 pandemic and mental health impacts. International journal of mental health nursing, 2020. 29(3): p. 315–318.

7. Kumar, A. and K.R. Nayar, COVID 19 and its mental health consequences. Journal of Mental Health, 2021. 30(1): p. 1–2.

8. Murata, S., et al., The psychiatric sequelae of the COVID-19 pandemic in adolescents, adults, and health care workers. Depression and Anxiety, 2021. 38(2): p. 233–246.

9. Estiri, H., et al., Evolving Phenotypes of non-hospitalized Patients that Indicate Long Covid. medRxiv, 2021: p. 2021.04.25.21255923.

10. Coppersmith, G., M. Dredze, and C. Harman. Quantifying mental health signals in Twitter. in Proceedings of the workshop on computational linguistics and clinical psychology: From linguistic signal to clinical reality. 2014.

11. McClellan, C., et al., Using social media to monitor mental health discussions ™ evidence from Twitter. Journal of the American Medical Informatics Association, 2017. 24(3): p. 496–502.

12. Yang, W. and L. Mu, GIS analysis of depression among Twitter users. Applied Geography, 2015. 60: p. 217–223.

13. Koh, J.X. and T.M. Liew, How loneliness is talked about in social media during COVID-19 pandemic: Text mining of 4,492 Twitter feeds. Journal of Psychiatric Research, 2020.

14. Valdez, D., et al., Social Media Insights Into US Mental Health During the COVID-19 Pandemic: Longitudinal Analysis of Twitter Data. J Med Internet Res, 2020. 22(12): p. e21418.

15. Gao, Y., Z. Xie, and D. Li, Electronic Cigarette Users’ Perspective on the COVID-19 Pandemic: Observational Study Using Twitter Data. JMIR Public Health and Surveillance, 2021. 7(1): p. e24859.

16. Hua, M., M. Alfi, and P. Talbot, Health-Related Effects Reported by Electronic Cigarette Users in Online Forums. J Med Internet Res, 2013. 15(4): p. e59.

17. Chen, L., et al., A Social Media Study on the Associations of Flavored Electronic Cigarettes With Health Symptoms: Observational Study. J Med Internet Res, 2020. 22(6): p. e17496.

18. Zou, C., et al., Public Reactions towards the COVID-19 Pandemic on Twitter in the United Kingdom and the United States. medRxiv: the preprint server for health sciences, 2020: p. 2020.07.25.20162024.

19. Gore, R.J., S. Diallo, and J. Padilla, You are what you tweet: connecting the geographic variation in america’s obesity rate to Twitter content. PloS one, 2015. 10(9): p. e0133505.

20. NLTK 3.6.2 documentation. Available from: https://www.nltk.org.

21. Gensim. Available from: https://radimrehurek.com/gensim_3.8.3/auto_examples/index.html.

22. Industrial-Strength Natural Language Processing. Available from: https://spacy.io.

23. Sievert, C. and K. Shirley. LDAvis: A method for visualizing and interpreting topics. in Proceedings of the workshop on interactive language learning, visualization, and interfaces. 2014.

24. Messias, J., P. Vikatos, and F. Benevenuto. White, man, and highly followed: Gender and race inequalities in Twitter. in Proceedings of the International Conference on Web Intelligence. 2017.

25. ethnicolr: Predict Race and Ethnicity From Name. Available from: https://ethnicolr.readthedocs.io/.

26. Sood, G. and S. Laohaprapanon, Predicting race and ethnicity from the sequence of characters in a name. arXiv preprint 1805.02109, 2018.

27. Auxier, B. and M. Anderson. Social Media Use in 2021. 2021; Available from: https://www.pewresearch.org/internet/2021/04/07/social-media-use-in-2021/.

28. Wang, C., et al., A longitudinal study on the mental health of general population during the COVID-19 epidemic in China. Brain, Behavior, and Immunity, 2020. 87: p. 40–48.

29. Jacques-Aviñó, C., et al., Gender-based approach on the social impact and mental health in Spain during COVID-19 lockdown: a cross-sectional study. BMJ Open, 2020. 10(11): p. e044617.

30. Guntuku, S.C., et al., Tracking Mental Health and Symptom Mentions on Twitter During COVID-19. Journal of General Internal Medicine, 2020. 35(9): p. 2798–2800.

31. Zhang, Y., et al., Monitoring depression trend on Twitter during the COVID-19 pandemic. arXiv preprint 2007.00228, 2020.

32. Lyu, H., et al., Sense and Sensibility: Characterizing Social Media Users Regarding the Use of Controversial Terms for COVID-19. IEEE Transactions on Big Data, 2020: p. 1–1.

